# Preexisting Comorbidities Predicting Severe Covid-19 in Older Adults in the UK Biobank Community Cohort

**DOI:** 10.1101/2020.05.06.20092700

**Authors:** Janice L Atkins, Jane AH Masoli, Joao Delgado, Luke C Pilling, Chia-Ling Kuo, George A Kuchel, David Melzer

**Affiliations:** Epidemiology and Public Health Group, University of Exeter Medical School, Exeter, UK; Department of Healthcare for Older People, Royal Devon and Exeter Hospital, Barrack Road, Exeter, UK.; Center on Aging, University of Connecticut Health Center, Farmington, CT, USA

**Keywords:** COVID-19, Morbidity, Epidemiology

## Abstract

**Background:** Older COVID-19 hospitalized patients frequently have hypertension, diabetes or coronary heart disease (CHD), but whether these are more common than in the population is unclear. During the initial epidemic in England, virus testing for older adults was restricted to symptomatic hospitalized patients. We aimed to estimate associations between pre-existing diagnoses and COVID-19 status, in a large community cohort.

**Methods:** UK Biobank (England) participants assessed 2006 to 2010, followed in hospital discharge records to 2017. Demographic and pre-existing common diagnoses association tested with COVID-19 status (16^th^ March to 14^th^ April 2020) in logistic models, adjusted for demographics, study site and other diagnoses.

**Results:** There were 274,356 participants aged 65+, including 448 (0.16%) hospitalized COVID-19 patients. Common co-morbidities in patients were hypertension (58.5%), coronary heart disease (CHD, 21.1%), history of fall or fragility fractures (30.6%), and type 2 diabetes (19.6%). However, in adjusted models, COVID-19 patients were more likely than other participants to have pre-existing dementia (OR=3.07 95% CI 1.71 to 5.50), COPD (OR= 1.82 CI 1.33 to 2.49), depression (OR=1.81 CI 1.36 to 2.40), type 2 diabetes (OR=1.70 CI 1.30 to 2.21), chronic kidney disease and atrial fibrillation. Hypertension was modestly associated (OR=1.29 CI 1.04 to 1.59), but CHD (OR=0.92 CI 0.71 to 1.20) prevalence was similar in COVID-19 patients and other participants.

**Conclusion:** Specific co-morbidities are disproportionally common in older adults who develop severe COVID-19. Tailored interventions may be needed, as these results do not support simple age-based targeting to prevent severe COVID-19 infection.

## Introduction

The 2019 novel coronavirus (SARS-COV-19) has caused a global pandemic [1], with a wide spectrum of clinical disease presentations, from asymptomatic infection to respiratory failure with high mortality [2]. However, little is known about what predicts COVID-19 severity in different individuals.

The majority of patients hospitalized with COVID-19 are older and have underlying medical conditions [3,4]. Analyses of COVID-19 patients have reported that increased age is associated with clinical severity [5,6], including case fatality [4,7]. The most frequent co-morbidities in Chinese cohorts of COVID-19 patients were hypertension (21.1%, 95% CI: 13.0–27.2%) and diabetes (9.7%, 95% CI: 7.2–12.2%), followed by cardiovascular disease (8.4%, 95% CI: 3.8–13.8%) and respiratory system disease (1.5%, 95% CI: 0.9–2.1%) [8] and in a large US cohort: hypertension (56.6%), obesity (41.7%), and diabetes (33.8%) [6]. However, co-morbidities vary with patient age-group, and in those aged over 70, obesity is less common and dementia is more common [3]. Such data on common co-morbidities in hospitalized patients is important for understanding the acute treatment challenge, but it is unclear whether these conditions are common in COVID-19 patients merely because they are also common in the older population. To identify those at most risk of severe COVID-19, data are needed on which pre-existing conditions are disproportionately common in severely affected patients compared to the background population.

The United Kingdom Biobank (UKB) is a community-based cohort of 500,000 participants currently aged 48 to 86 [9]. UKB recently added linkage to National Health Service COVID-19 laboratory test results in England for the period March 16 to April 14 2020, thus including the peak period of daily COVID-19 laboratory-confirmed cases in the current outbreak [10]. During this period, testing of older groups was essentially restricted to hospital in-patients with clinical signs of infection [11], so test positivity is a good marker of severe COVID-19 [12]. Given the scarcity of cohort data on risk factors for severe COVID in older groups, we undertook an initial analysis of baseline (2006 to 2010) demographic characteristics and pre-existing diagnoses during UK Biobank (UKB) follow-up. Identifying the specific risk factors explaining why some older people are severely affected by COVID-19 than others may give clues to underlying vulnerabilities, and are critical to developing outbreak control policies that focus on individual risk and avoid imposing crude age based public health controls [13].

## Methods

### UK Biobank Cohort

The UK Biobank consists of over 500,000 community volunteers aged 40 to 70 year at baseline (2006 to 2010), living close to 22 assessment centers in England, Scotland and Wales [9]. Baseline assessments included demographics, lifestyle and disease history, with linkages to electronic medical records. UK Biobank ethical approval was from the North West Multi-Centre Research Ethics Committee. The current analysis was approved under UKB application 14631 (PI Melzer, D).

### COVID-19 test status and sample selection

Of 1,474 UK Biobank participants of all ages with available data, 669 were laboratory-confirmed active COVID-19 infection (by PCR) from test preformed between March 16 and April 14, 2020, in England only. During this period, COVID-19 testing was restricted to hospitalized patients with clinical signs of the disease, plus health care workers [14, 15]. We restricted analyses to participants aged 65 years and older when tested (or age on April 14 if not tested), to minimize misclassification of disease severity from health care workers (who were tested even for mild possible infections): this included 448 older study participants testing COVID-19 positive. We also excluded participants previously reported to have died during UK Biobank follow-up (n=20,348, death registration records available until January 2018). No COVID-19 test data were available for UKB assessment centers in Scotland and Wales, so data from these centers were not included.

### Disease ascertainment

Preexisting diagnoses were available from baseline questionnaires (2006-10) eliciting participant reports of doctor diagnosed disease. New disease diagnoses since baseline were from linked electronic medical records to hospital inpatient routine data (to March 2017), coded according to the International Classification of Diseases 10^th^ revision (ICD-10). Diagnoses included were coronary heart disease (CHD), atrial fibrillation, stroke, hypertension, diabetes (type 2), chronic kidney disease (CKD, stages 3 to 5), depression, dementia, asthma, chronic obstructive pulmonary disease (COPD), osteoporosis and osteoarthritis. We also identified previous diagnoses of delirium, pneumonia and falls or fragility fractures (See definitions in Supplementary Table 1). We combined each diagnosis reported at baseline or from linked hospital data to generate pre-existing diagnosis status for each participant.

### Statistical Analysis

We estimated associations of demographic and diagnoses with COVID-19 test positivity using logistic regression models, with 95% confidence intervals. Logistic models were adjusted for age group (in five year age bands), sex, ethnicity, education and assessment center at baseline (to account for geographic differences in the prevalence of COVID-19 infection). A p-value smaller than 5% was considered statistically significant. All the statistical analyses were performed in Stata version 15.1.

## Results

There were 274,356 older adults (aged 65 to 86 years, mean 73.2 years) eligible for the analysis, of whom 448 (0.16%) were laboratory test positive COVID-19 patients, with rates of infection varying widely across UK Biobank baseline assessment centers (0.38% to 0.07%, see Supplementary Table 2). The mean age of patients (Table 1) was 74.3 years (SD 4.5) in COVID-19 test positive patients and 73.2 years (SD 4.4) for other study participants. Both patients and others were predominantly of ‘white’ ethnicity, but people self-reporting ‘Black’ ethnicity made up 5.2% of patients but only 1% of participants. The most common diagnoses in cases were hypertension (58.5%), a history of falls or fragility fractures (30.6%), CHD (21.1%), diabetes (type 2, 19.6%), asthma (18.2%). Dementia was present in 3.4% of patients (0.5% of other participants). COVID-19 patients had a mean of 2.4 diagnoses (of a possible 15 diagnoses examined) compared to other participants with 1.4 diagnoses.

**Table 1.**
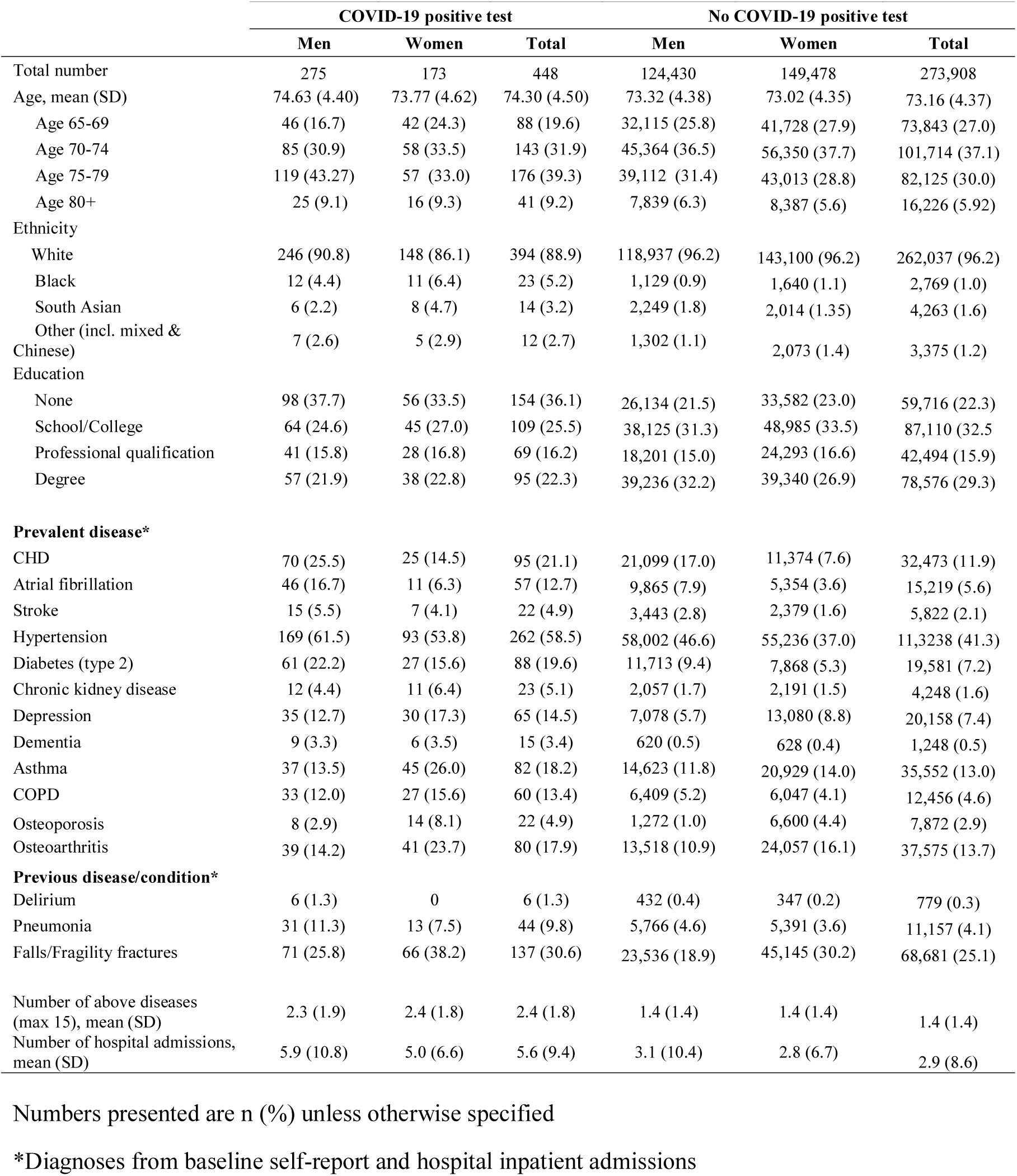
Descriptive characteristics of UK Biobank cohort by COVID-19 test positivity

In logistic modelling of demographic variables (Table 2, ‘demographics’, also accounting for assessment center), risks in those aged 80+ years were markedly raised (Odds Ratio OR=2.24 95% CI 1.53 to 3.30, p=4.10E-05) compared to age-group 65 to 69, with no risk increase in 70 to 74 years old and intermediate risks in 75 to 80 years old. Males were substantially more likely to be COVID-19 test positive patients (OR=1.89 CI 1.56 to 2.30, p=1.60E-10) and people of Black ethnicity were at higher risk (OR 4.05, CI 2.55 to 6.43, p=2.90E-09) compared to White, with South Asians and other ethnicities having intermediate risks. Compared to those with degree level education, having no education qualifications (OR 2.10 CI 1.61 to 2.74, p=4.60E-08) was associated with raised risks of COVID-19. All studied diagnoses individually (adjusted for demographics only) were associated with COVID-19 (Supplementary Table 3). As different chronic diseases often co-exist together in older adults, we estimated risks for each diagnosis accounting for other diagnoses present. In fully adjusted models (Table 2, Full model), the excess risk in men was virtually unchanged by adjustment for diagnoses (OR 1.83 (1.49 to 2.25, p=7.00E-09), but there were somewhat lower risks in the 80+ year olds (OR 1.76 CI 1.19 to 2.62, p=5.00E-03, compared to 65 to 69). Also, dementia emerged as being associated with the largest increase in risks of COVID-19 (OR=3.07 CI 1.71 to 5.50, p=1.60E-04) followed by COPD (OR 1.82 CI 1.33 to 2.49, p=1.90E-04), depression (OR 1.81 CI 1.36 to 2.4, p=3.90E-05), diabetes (OR=1.7 CI 1.3 to 2.21, p=8.90E-05), chronic kidney disease (OR 1.60 CI 1.03 to 2.49, p=3.80E-02) and atrial fibrillation (OR 1.51 CI 1.12 to 2.05, p=7.70E-03), with a relatively modest risk increase with hypertension (OR= 1.29 CI 1.04 to 1.59, p=2.00E-02). Coronary heart disease (CHD) prevalence, previously noted as common in COVID-19 patients, did not differ between patients and other participants (OR=0.92 CI 0.71 to 1.20, p=5.30E-01) in the models adjusted for demographics and other diagnoses. In sex specific analyses (Table 3) of COVID-19, there were associations in women for dementia (OR=5.22 CI 2.21 to 12.31, p=1.60E-04), depression, chronic kidney disease, asthma and COPD, and in men, dementia, depression, atrial fibrillation and diabetes reached statistical significance: however, the confidence intervals overlapped for the diagnoses that differed by sex.

**Table 2.** Odds ratios for associations between A) demographic variables and COVID-19 status, and B) pre-existing diagnoses and COVID-19, accounting for demographics plus other diagnoses (full adjustment model)

|  | Demographics |  | Full model |  |
| --- | --- | --- | --- | --- |
|  | OR (95% CI) | p-value | OR (95% CI) | p-value |
| Age |  |  |  |  |
| Age 65-69 |  |  |  |  |
| Age 70-74 | 1.16 (0.88 to 1.53) | 2.90E-01 | 1.11 (0.84 to 1.46) | 4.80E-01 |
| Age 75-79 | 1.65 (1.26 to 2.16) | 2.60E-04 | 1.44 (1.09 to 1.89) | 9.20E-03 |
| Age 80+ | 2.24 (1.53 to 3.30) | 4.10E-05 | 1.76 (1.19 to 2.62) | 5.00E-03 |
| Sex |  |  |  |  |
| Female |  |  |  |  |
| Male | 1.89 (1.56 to 2.30) | 1.60E-10 | 1.83 (1.49 to 2.25) | 7.00E-09 |
| Ethnicity |  |  |  |  |
| White |  |  |  |  |
| Black | 4.05 (2.55 to 6.43) | 2.90E-09 | 3.69 (2.31 to 5.89) | 4.60E-08 |
| South Asian | 1.82 (1.03 to 3.22) | 3.90E-02 | 1.54 (0.87 to 2.75) | 1.40E-01 |
| Other (inc. mixed & Chinese) | 1.98 (1.05 to 3.75) | 3.60E-02 | 1.81 (0.95 to 3.44) | 6.90E-02 |
| Education |  |  |  |  |
| Degree |  |  |  |  |
| Professional qualification | 1.42 (1.04 to 1.95) | 2.80E-02 | 1.33 (0.97 to 1.83) | 7.40E-02 |
| School/College | 1.12 (0.85 to 1.48) | 4.30E-01 | 1.04 (0.78 to 1.37) | 8.10E-01 |
| None | 2.10 (1.61 to 2.74) | 4.60E-08 | 1.71 (1.30 to 2.24) | 1.10E-04 |
| **Prevalent disease\*** |  |  |  |  |
| Coronary heart disease |  |  | 0.92 (0.71 to 1.20) | 5.30E-01 |
| Atrial fibrillation |  |  | 1.51 (1.12 to 2.05) | 7.70E-03 |
| Stroke |  |  | 1.25 (0.79 to 1.96) | 3.40E-01 |
| Hypertension |  |  | 1.29 (1.04 to 1.59) | 2.00E-02 |
| Diabetes (type 2) |  |  | 1.70 (1.3 to 2.21) | 8.90E-05 |
| Chronic kidney disease |  |  | 1.60 (1.03 to 2.49) | 3.80E-02 |
| Depression |  |  | 1.81 (1.36 to 2.4) | 3.90E-05 |
| Dementia |  |  | 3.07 (1.71 to 5.5) | 1.60E-04 |
| Asthma |  |  | 1.13 (0.87 to 1.47) | 3.60E-01 |
| COPD |  |  | 1.82 (1.33 to 2.49) | 1.90E-04 |
| Osteoporosis |  |  | 1.42 (0.91 to 2.23) | 1.20E-01 |
| Osteoarthritis |  |  | 1.10 (0.85 to 1.42) | 4.90E-01 |
| **Previous disease/condition\*** |  |  |  |  |
| Delirium |  |  | 1.22 (0.51 to 2.90) | 6.60E-01 |
| Pneumonia |  |  | 1.40 (0.99 to 1.97) | 5.50E-02 |
| Falls/Fragility fractures |  |  | 1.15 (0.92 to 1.42) | 2.10E-01 |

\*Diagnoses from baseline self-report and hospital inpatient admissions
Demographics model (adjusted for age group, sex, ethnicity, education, and baseline assessment centre)
Full model (adjusted for age group, sex, ethnicity, education, assessment centre and all the above diseases/conditions)

**Table 3.** Odds ratios for associations between pre-existing diagnoses and COVID-19 status, accounting for demographics plus other diagnoses (full adjustment model), stratified by sex

|  | WOMEN |  | MEN |  |
| --- | --- | --- | --- | --- |
|  | Full adjustment |  | Full adjustment |  |
|  | OR (95% CI) | p-value | OR (95% CI) | p-value |
| **Prevalent disease\*** |  |  |  |  |
| Coronary heart disease | 0.82 (0.50 to 1.36) | 4.50E-01 | 0.96 (0.70 to 1.31) | 7.90E-01 |
| Atrial fibrillation | 1.06 (0.54 to 2.07) | 8.70E-01 | 1.71 (1.21 to 2.41) | 2.40E-03 |
| Stroke | 1.29 (0.56 to 3.00) | 5.50E-01 | 1.22 (0.71 to 2.10) | 4.60E-01 |
| Hypertension | 1.36 (0.98 to 1.9) | 7.00E-02 | 1.24 (0.94 to 1.63) | 1.30E-01 |
| Diabetes (type 2) | 1.6 (1.00 to 2.55) | 5.00E-02 | 1.76 (1.27 to 2.42) | 5.80E-04 |
| Chronic kidney disease | 2.29 (1.19 to 4.38) | 1.30E-02 | 1.24 (0.67 to 2.28) | 4.90E-01 |
| Depression | 1.69 (1.11 to 2.58) | 1.40E-02 | 1.90 (1.30 to 2.78) | 9.60E-04 |
| Dementia | 5.22 (2.21 to 12.31) | 1.60E-04 | 2.33 (1.07 to 5.05) | 3.20E-02 |
| Asthma | 1.47 (1.01 to 2.14) | 4.40E-02 | 0.90 (0.62 to 1.31) | 5.80E-01 |
| COPD | 2.59 (1.60 to 4.18) | 1.00E-04 | 1.49 (0.98 to 2.25) | 6.00E-02 |
| Osteoporosis | 1.21 (0.68 to 2.15) | 5.20E-01 | 1.90 (0.92 to 3.91) | 8.30E-02 |
| Osteoarthritis | 1.17 (0.81 to 1.70) | 4.10E-01 | 1.01 (0.70 to 1.44) | 9.80E-01 |
| **Previous disease/condition\*** |  |  |  |  |
| Delirium | No observations |  | 2.08 (0.86 to 5.04) | 1.10E-01 |
| Pneumonia | 1.25 (0.68 to 2.30) | 4.60E-01 | 1.48 (0.98 to 2.26) | 6.50E-02 |
| Falls/Fragility fractures | 1.15 (0.84 to 1.59) | 3.90E-01 | 1.14 (0.86 to 1.52) | 3.70E-01 |

\*Diagnoses from baseline self-report and hospital inpatient admissions
Full model (adjusted for age group, ethnicity, education, assessment centre and all the above diseases/conditions).

## Discussion

We aimed to identify risk factors associated with COVID-19 test positivity in older hospital inpatients during the peak of the initial epidemic in England. We found that some chronic conditions reported as common co-morbidities in COVID-19 patients (and postulated as major risk factors), were modestly (diabetes, hypertension) or not (CHD) significantly more common with COVID-19 than in other older UK Biobank participants. Instead, pre-existing diagnoses of dementia, COPD, depression, chronic kidney disease and atrial fibrillation emerged as independent risk factors. In keeping with other studies, we also found increased risk of COVID-19 in males, but we have shown that this risk is virtually unchanged after adjusting for co-morbidities. In addition, we confirmed previous reports that people of Black ethnicity, and those with no educational qualifications had higher risk. Overall, these results suggest that there are specific risk co-morbidities in older groups, and that severe COVID-19 susceptibility is not merely the result of advancing age.

Hypertension is well-recognized as the most common chronic diagnosis, prevalent in over 70% of persons at ages over 80 [16]. While highly prevalent in the UKB cohort, we found that it was only modestly more common in COVID-19 cases than in other UKB participants. Interestingly, we found that having a diagnosis of CKD (grade 3 to 5) was associated with being COVID-19 positive, mirroring previous analyses by Masoli et al [17], who found that CKD grade is more predictive of mortality than blood pressure in adults age over 70. CKD has also been reported to be associated with increased hospitalisation with infection, particularly pneumonia, and increased 30 day mortality [18, 19].

Another novel finding is the association between atrial fibrillation and hospitalized COVID-19 in the studied older adults. During atrial fibrillation, the loss of atrio-ventricular synchrony with decreased diastolic filling time is likely to lead to a decrease in cardiac output. Consequently, this low cardiac output may aggravate tissue hypoxia in COVID-19 patients. Also, agents used in the control of atrial fibrillation, particularly sotalol, propafenone, and non-selective β-blockers, may cause bronchospasm [20]. Pulmonary symptoms in COPD may become worse with atrial fibrillation development, due to excessive irregular heart rate, as well as reduced diastolic filling of the ventricles [21]. These factors may contribute to higher severe COVID-19 risk in participants with atrial fibrillation.

The relationship between diabetes and COVID-19 has been widely noted, with diabetes associated with increased severity [22]. However, the association between diabetes and clinical severity of COVID-19 in diabetes reduced at older ages, compared to under 55 year olds [23], and we found a relatively modest association between diabetes and COVID-19 in our study. Unfortunately, the UK Biobank data lacks a recent measure of adiposity, but the effects of obesity are likely evident in the raised risks with diabetes and other related conditions.

To date there has been very limited data on dementia and COVID-19, despite dementia affecting over 50 million people worldwide [24]. This may be due to the young median ages of many published COVID-19 case series, with limited characterisation of older persons. Our analysis has revealed dementia to be the largest effect risk diagnosis in adults aged over 65, in this cohort of community volunteers. A recent report of observational data from the International Severe Acute Respiratory and Emerging Infections Consortium (ISARIC) found a high prevalence of dementia in older adults admitted to hospital with COVID-19 [3]. Future work will need to clarify the mechanisms involved and aim to establish whether this a direct effect of dementia pathologies, or an indirect effect of high rates of infection in nursing homes.

UK Biobank participants were somewhat healthier than the general population [25] at baseline in 2006 to 2010, but the sample nevertheless includes large numbers of socio-economically less privileged participants: for example, 36.1% of cases and 22.3% of controls had no educational qualifications (Table 1). Other limitations include the lack of details of the hospitalization and degrees of clinical severity within cases, although at the peak of the epidemic, access to intensive care units for older patients may partly reflect resource scarcity rather than severity of illness. Only a small proportion of the English population has thus far been exposed to the virus, but the group studied here were exposed and developed severe enough COVID-19 to be tested during hospitalization. Our case group is therefore relevant for assessing risk factors for severe COVID-19 in this older population. Our diagnostic data is derived from participant’s baseline interviews plus hospital discharge data until March 2017, so under-ascertainment of disease is likely, especially for recently diagnosed conditions, but the similarity to previous reports of the common conditions seen in COVID-19 patients suggests that our data are valid.

Our results should have implications for preventive interventions, encouraging a more targeted approach prioritizing those older adults with specific risk factors, rather than adopting policies that use chronological age as a blanket indicator of risk. Our cohort evidence of specific risk factors may also help with avoiding potentially ‘ageist’ approaches to setting clinical priorities in over-stretched health systems [26]. Our findings of risks associated with less prominent conditions such as atrial fibrillation and CKD could help focus clinical research, as patients these conditions may need specific interventions to reduce risks of COVID-19 severity. In addition, the prominence of depression as one of the major risk factors highlights the role of mental health [27] as critical to managing the pandemic, including in older people.

## Conclusion

In older adults, several specific pre-existing co-morbidities are disproportionally common in severe COVID-19 patients, notably including dementia, depression, atrial fibrillation and chronic kidney disease. Clinical and public health research is needed to establish the mechanisms involved and whether stratified interventions are needed for older patients with specific comorbidities. A history of coronary heart disease is common in both COVID-19 hospitalized patients and other study participants. Our results do not support simple age targeting of interventions to prevent severe COVID-19 infection.

## Data Availability

Data are available on application to the UK Biobank (www.ukbiobank.ac.uk/register-apply).

## Acknowledgements

This research was conducted using the UK Biobank resource, under application 14631. We thank the UK Biobank participants and coordinators for the dataset.

## Author contributions

JLA & DM performed the statistical analysis of data. DM, JAHM, JD and JLA drafted the manuscript. All authors were involved in design of the study, interpretation of data and revision of the manuscript.

